# Catheter Ablation Compared to Medical Therapy for Ventricular Tachycardia in Sarcoidosis: Outcomes and Hospital Readmissions in a Nationwide Cohort Study

**DOI:** 10.1101/2023.05.05.23289599

**Authors:** Michael I. Gurin, Yuhe Xia, Constantine Tarabanis, Randal I. Goldberg, Robert J. Knotts, Alex Reyentovich, Robert Donnino, Scott Bernstein, Lior Jankelson, Alexander Kushnir, Douglas Holmes, Michael Spinelli, David S. Park, Chirag R. Barbhaiya, Larry A. Chinitz, Anthony Aizer

## Abstract

**Background:** Patients with cardiovascular manifestations of sarcoidosis are at an increased risk for ventricular arrhythmias (VA) and sudden cardiac death. Catheter ablation (CA) for ventricular tachycardia (VT) can be a useful treatment strategy, however, few studies have compared CA to medical therapy in this patient population.

**Objective:** To assess in-hospital outcomes and unplanned readmissions following CA for VT compared to medical therapy in patients with sarcoidosis.

**Methods:** Using ICD-9 and ICD-10 diagnostic and procedural codes, data was obtained from the Nationwide Readmissions Database between January 2010 and December 2019 to identify patients with a diagnosis of sarcoidosis admitted for VT either undergoing CA or medical therapy. Primary endpoints were 30-day unplanned hospital readmissions as well as a composite endpoint of inpatient mortality, cardiogenic shock, and cardiac arrest. Complications at index hospitalization and causes of readmission were also identified.

**Results:** Among a total of 1,581 patients, 1,349 patients with sarcoidosis and a diagnosis of VT were managed medically compared to 232 that underwent CA. Readmission rates at 30 days were 10.8% and 8.0%, respectively (*p*=0.266). In univariate analysis, the composite endpoint of mortality, cardiac arrest and cardiogenic shock trended in favor of ablation (7.4% vs 11.7%, *p*=0.067). In the subgroup of patients undergoing elective CA for VT, there was an improvement in the univariate composite of mortality, cardiac arrest, and cardiogenic shock (3.2% vs. 11.7%, *p*=0.039). After multivariable adjustment, patients undergoing elective CA were less likely to be readmitted within 30-days (OR 0.23 [95% CI 0.05,0.90] *p*=0.042). The most common cause of readmission were VA in both groups, however, those undergoing elective CA were less likely to be readmitted for VA compared to non-elective ablation. Complications in the CA group included cardiac tamponade (4.7%), vascular complications (2.6%), and hematomas (3.0%).

**Conclusion:** In a national database of patients admitted with sarcoidosis and VT, when compared to medical therapy, CA results in a similar 30-day readmission rate with a trend towards reduction in the univariate composite endpoint of inpatient mortality, cardiogenic shock, and cardiac arrest.

Patients undergoing elective VT ablation have a superior univariate outcome in the primary composite endpoint and were less likely to be readmitted within 30-days in adjusted analysis compared to medical therapy. Procedure related complications were low in the ablation group. The findings of short-term safety compared to medical therapy in addition to early intervention adds further support to an elective CA approach.

**Clinical Perspective What is New?:**

- We report nationwide in-hospital outcomes and readmission rates in sarcoidosis patients presenting with ventricular tachycardia (VT) undergoing catheter ablation (CA) as compared to medical therapy alone.
- Elective catheter ablation shows a superior reduction in a composite endpoint of inpatient mortality, cardiogenic shock, and cardiac arrest and are less likely to be readmitted within 30-days compared to medical therapy.
- Ventricular arrhythmias (VA) are the most common cause of readmission, however, patients undergoing elective CA are less likely to be readmitted for VA.

**What Are the Clinical Implications?:**

- VT ablation in sarcoidosis patients favors an elective ablation strategy over medical therapy alone, making pre-procedural optimization, patient selection, and timing critical for successful catheter ablation.
- Provides clinicians with guidance in formulating acute management decisions in sarcoid patients presenting with VT.
- Patients undergoing unplanned CA for VT as compared to elective CA have similar complication rates and no obvious increased risk of harm, suggesting that CA can be an important bailout for patients who cannot afford to wait until an elective ablation is performed.

## Introduction

Patients with cardiovascular manifestations of sarcoidosis are at increased risk for ventricular arrhythmias (VA) and sudden cardiac death. Estimates of 5-year mortality can exceed 40%^1^ with VA and congestive heart failure being leading predictors of morbidity and mortality.^2–4^ The complex arrhythmic substrate in cardiac sarcoidosis (CS) can be further affected by an underlying inflammatory state, making management of ventricular tachycardia (VT) all the more challenging.^5, 6^ Immunosuppression, antiarrhythmic medications and implantable cardioverter defibrillators (ICDs) have been shown to be a feasible treatment strategy for VT in CS^7^, with catheter ablation (CA) emerging in recent decades as an option for drug-refractory VT.^8, 9^

Because of the unique substrate and varied outcomes of sarcoidosis VT patients, determining optimal treatment strategies remains elusive. Current CS expert consensus recommends a stepwise immunosuppression and antiarrhythmic therapy for VA with evidence of myocardial inflammation, with catheter ablation reserved for VA refractory to medical therapy.^10^ Although catheter ablation can be helpful in a number of cases in CS, VT recurrence rates are high, and among non-ischemic cardiomyopathies, patients with CS have the worst prognosis with respect to all-cause death, need for heart transplantation, and hospital readmission.^11–13^ In contrast to this, there is emerging data that in other patient populations, early, optimized VT ablation may have more favorable long term outcomes.^14–16^ To our knowledge, limited studies have compared acute or long term outcomes between CS patients undergoing nonelective VT ablation, inpatient medical therapy or elective VT ablation. As a result, there is little data to guide management in the acute setting.

The primary objective of this study is to assess in-hospital outcomes and unplanned readmissions following catheter ablation for VT in patients with sarcoidosis as compared to those with sarcoidosis admitted for VT undergoing medical therapy alone. Additionally, we compared in-hospital outcomes and readmission rates in both elective and unplanned (non-elective) hospital admissions in patients undergoing catheter ablation for VT for cardiac sarcoidosis.

## Methods

### Data source

Data was obtained from the Nationwide Readmissions Database (NRD) between January 1, 2010 and December 31, 2019. The NRD is a publicly available deidentified database of hospitalizations in the United States specifically designed for readmission analyses and sponsored by the Agency for Healthcare Research and Quality (AHRQ) as part of the Healthcare Cost and Utilization Project. The database contains more than 100 clinical and nonclinical variables for each hospital stay, including diagnosis and procedure codes from the International Classification of Disease, Ninth and Tenth Revisions, Clinical Modification (ICD-9-CM and ICD-10-CM), and data from all payers as well as uninsured persons. Discharge data is available from 30 geographically dispersed states, accounting for 61.8% of the total U.S. population and 60.4% of all U.S. hospitalizations. As the NRD is a publicly available deidentified database, this study did not qualify as human subject research and is exempt from institutional review board approval.

### Study population

The ICD-9-CM and ICD-10-CM codes specified in Supplemental Table 1 were used to identify patients aged 18 and older from the NRD with a diagnosis of sarcoidosis undergoing catheter ablation or sarcoidosis without catheter ablation. Patients were further selected by having a primary hospital admission for VT. Admissions with a secondary diagnosis of atrial fibrillation, supraventricular tachycardia, and atrial flutter were excluded due to overlap in the billing code used for atrial and ventricular ablations. The NRD also contains data elements that permit further sub-selection of patients based on elective versus non-elective admission. The hospitalization during which VT catheter ablation or admission for VT took place was defined as the index admission (Figure 1). To mitigate the risk of incomplete follow up, hospitalizations occurring in a state other than a patient’s primary residence or during the month of December of each year did not contribute to admissions. The latter allowed all patients to have uniform follow-up of 30 days. Subsequently, admissions were excluded between September and December for 90-day analysis and from July to December for 180-day analysis. Any planned readmission and patients who died during the index admission were excluded from the readmission analysis.

**Figure 1.**
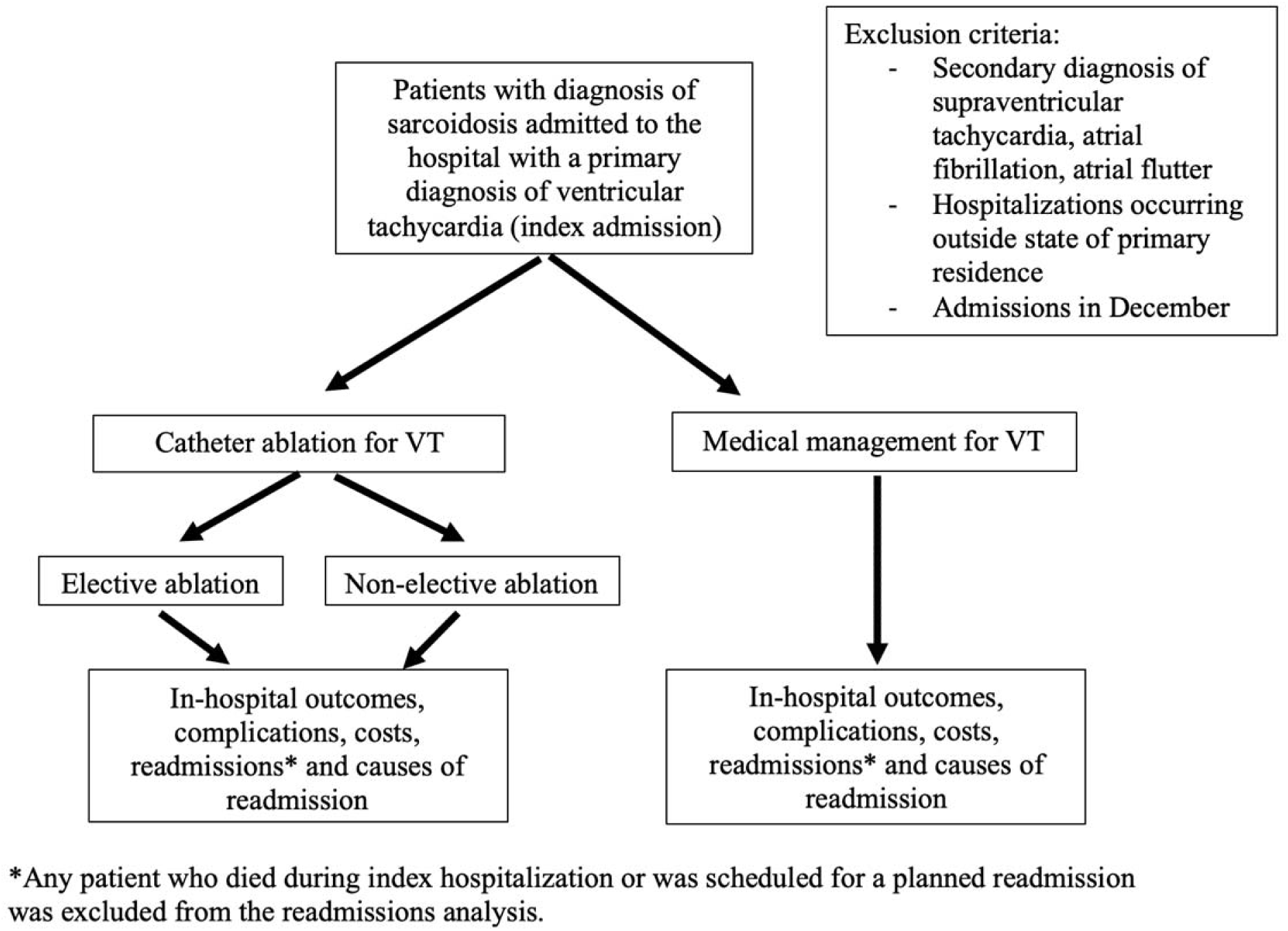
Patient selection, inclusion and exclusion criteria.

### Study endpoints

The two primary study endpoints were unplanned all-cause 30-day readmissions and a composite endpoint of mortality, cardiogenic shock, and cardiac arrest. Secondary endpoints were 90 and 180-day readmission rates, rates of mechanical circulatory support, cardiogenic shock, and cardiac arrest. Complications and causes of readmission were identified using the ICD codes available in the Supplemental Table 1. Causes of readmission were reported, stratified by cohort.

### Statistical analysis

Demographics, comorbidities, and outcomes were compared between the cohorts using chi-square analysis or Fisher’s exact tests for categorical variables, and Student’s t-test or Mann-Whitney U test for continuous variables. A two-sided p<0.05 was considered to be statistically significant. Covariates were used for multivariable logistic regression if after univariate analysis they were significant at p<0.1 and if the covariate was clinically relevant to the outcome variable. We performed two multivariable regressions, one to compare catheter ablation to medical therapy, and a second multivariable regression to compare elective to non-elective catheter ablation. In the first multivariable regression, we adjusted for the following independent variables: elective admission, age, gender, heart failure, atrioventricular block, left bundle branch block, immunosuppression/steroid use, hypertension, chronic obstructive pulmonary disease, diabetes mellitus, renal failure, liver disease, fluid and electrolyte disorders, coagulopathy, anemia, obesity, hospital bed size, teaching status (teaching vs. non-teaching hospitals), size of metropolitan area, and presence of pacemaker or implantable cardioverter-defibrillator. Non-metropolitan hospitals were removed from the analysis due to limited sample size. The second multivariable regression analysis was performed to compare elective and non-elective ablation. For this analysis, chronic obstructive pulmonary disease, heart failure, liver disease, coagulopathy, and fluid and electrolyte disorders were removed from the model due to being in the causal pathway between the exposure and outcomes. Multivariate regression was used to estimate the odds of readmission, complications, mortality, and any adverse outcome (cardiac arrest, cardiogenic shock, and mortality) after adjusting for the differences in demographics and comorbidities between the cohorts. All analyses were conducted using Rstudio Version 4.0.3.

## Results

### Baseline characteristics

During 2010-2019, there were 1581 patients meeting study criteria. Of these, 1349 patients had sarcoidosis and VT managed medically. The patients managed medically were mostly male (60.5%) and the average age was 56 years. Primary insurance was private (55.3%) followed by Medicare (34.8%) and patients were represented across all quartiles of median household income. Sarcoidosis patients clustered in areas with larger populations including metropolitan areas where the population was greater than 1 million persons and in nearby counties to metropolitan areas with greater than 1 million persons. There were 232 patients meeting inclusion criteria for sarcoidosis undergoing CA for VT. These were also mostly male (70.7%) with an average age of 54 years. Private insurance was most common (62.1%) and patients were also distributed evenly among all quartiles of median household income with those in the highest income bracket predominating (34.5%). Similarly, most patients were in large metropolitan areas and fringe counties in metropolitan areas of greater than 1 million persons. Sarcoidosis patients presented more frequently to large sized hospitals, were affiliated with a metropolitan teaching hospital (84.5%), and were found in large metropolitan areas (Table 1).

**Table 1.**
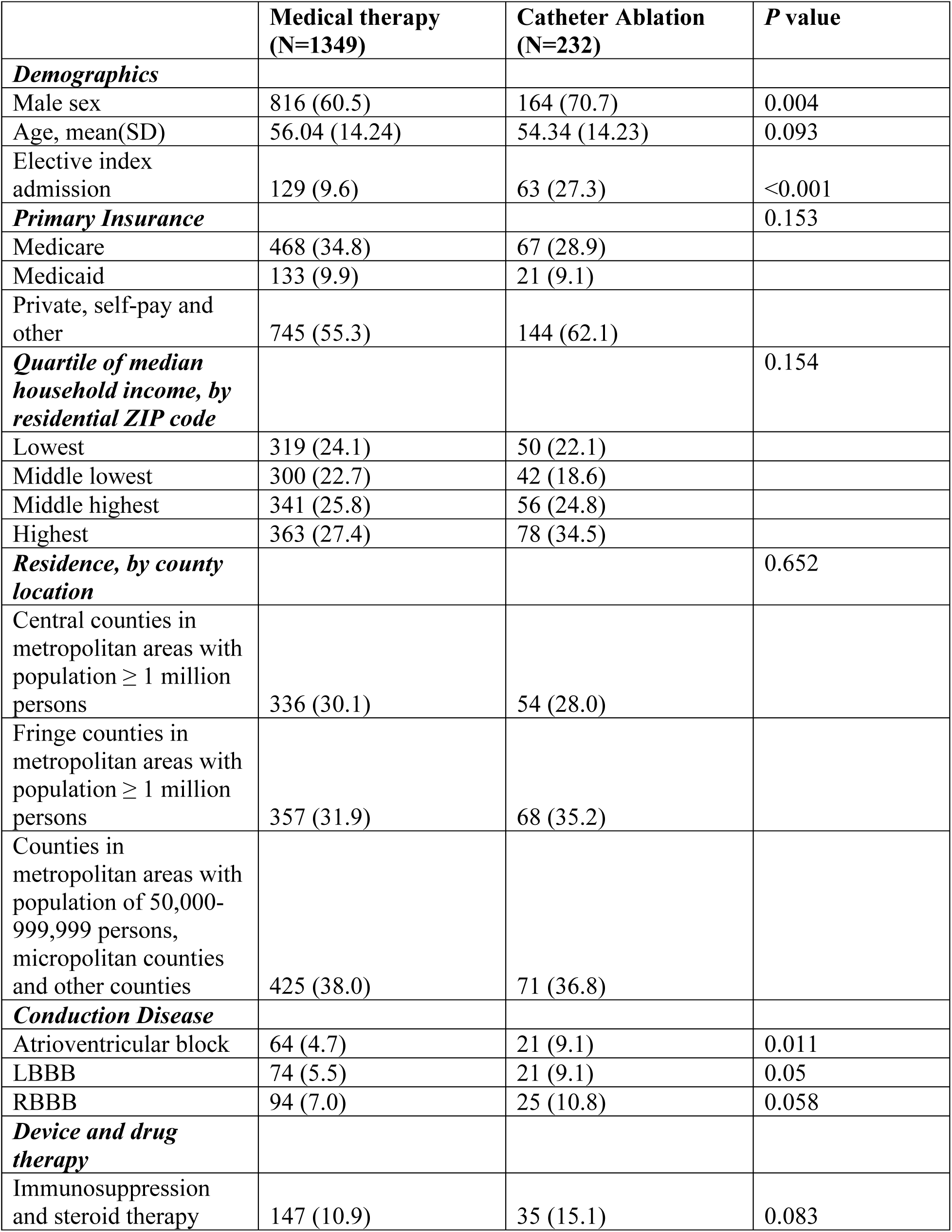
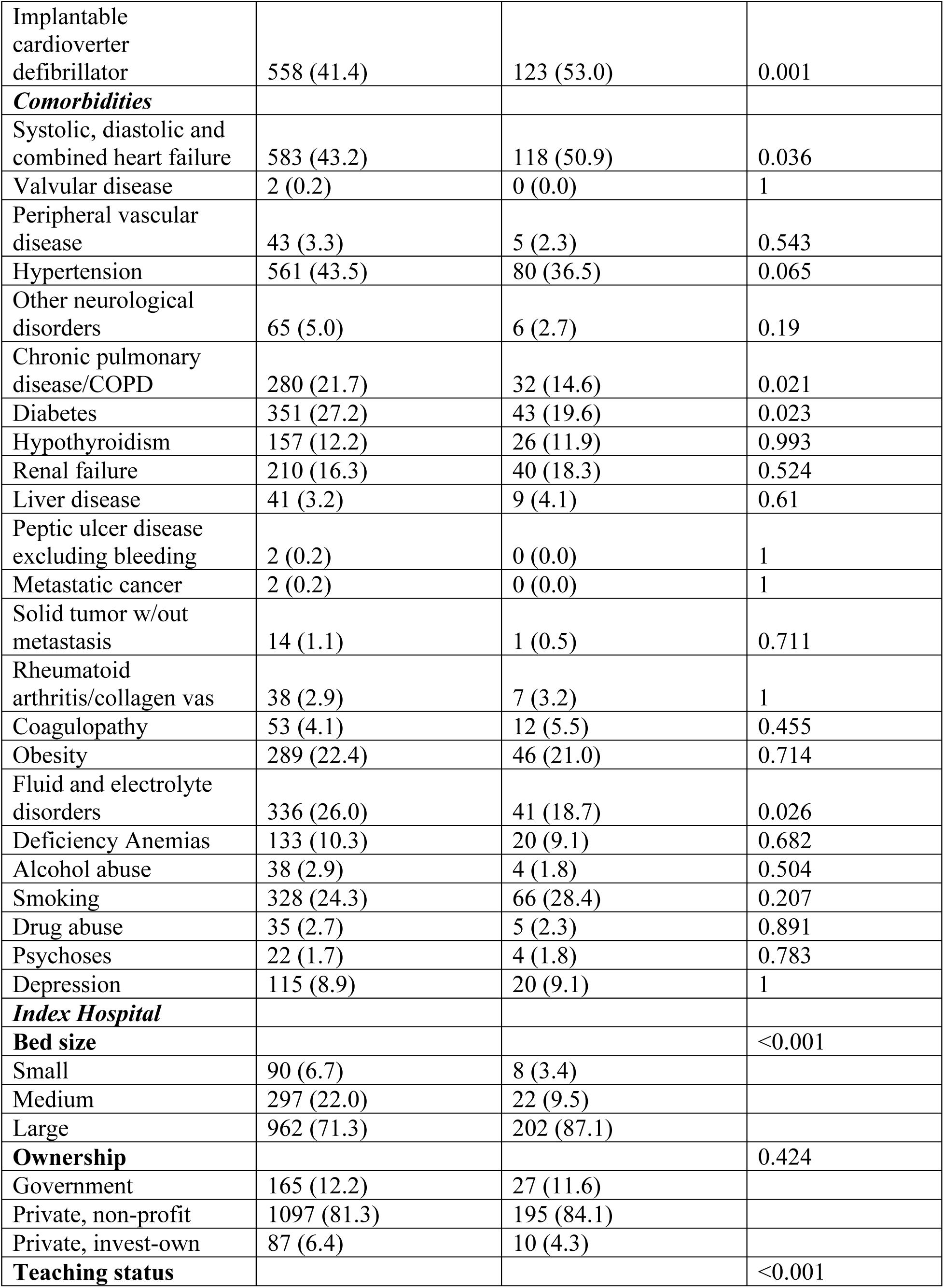
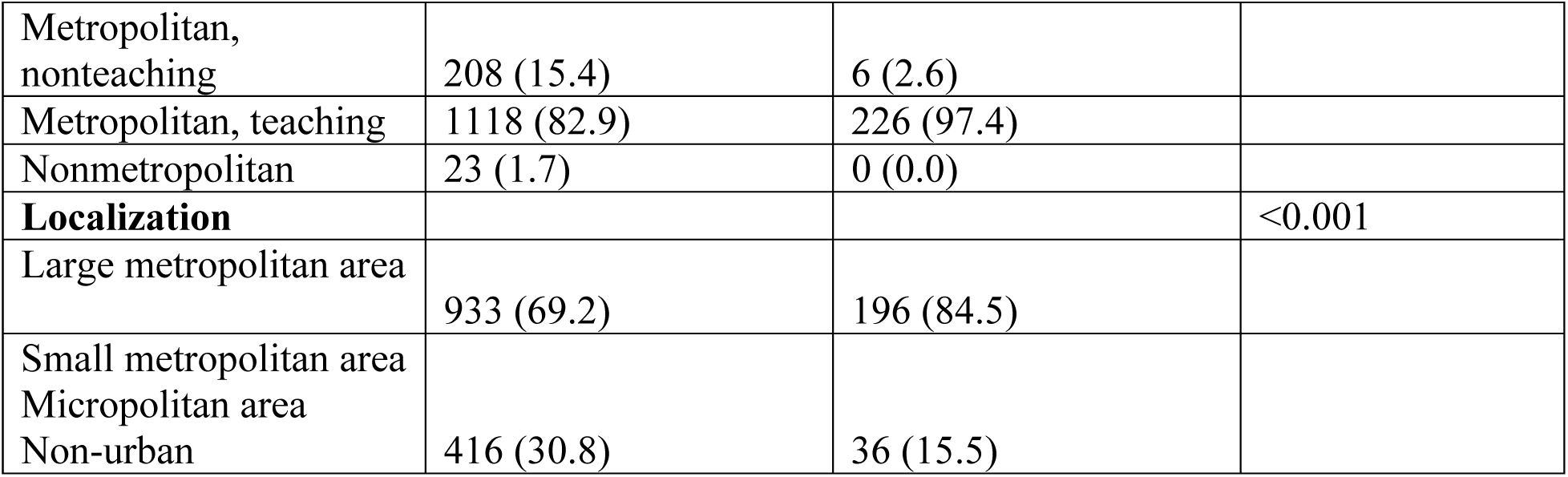
Baseline characteristics, sarcoidosis patients managed medically versus catheter ablation. Demographic and baseline health patient characteristics, along with the institutional characteristics corresponding to patients with sarcoidosis managed medically versus with catheter ablation, stratified by cohort. SD=standard deviation, LBBB=left bundle branch block, RBBB=right bundle branch block, COPD=chronic obstructive pulmonary disease, IQR=interquartile range

At index hospitalization for VT ablation, about one quarter of patients were admitted electively (27.3%) compared with 9.6% elective admission rate for medical therapy. Compared to sarcoidosis patients medically managed, patients undergoing VT CA had greater prevalence of heart failure (50.9% vs. 43.2%), conduction disease including left (9.1 vs. 5.5%) and right bundle branch block (10.8% vs. 7.0%), as well as atrioventricular block (9.1% vs. 4.7%), and an implantable cardioverter-defibrillator (53.0% vs 41.4%). There was no statistically significant difference between groups in history of or current use of immunosuppression and steroid therapy. In patients medically managed for VT, there was greater prevalence of chronic pulmonary disease (21.7% vs 14.6%), diabetes (27.2% vs 19.6%), and fluid and electrolyte disorders (26.0 vs 18.7%) (Table 1).

### In-hospital outcomes

Compared to medical therapy, in-hospital complications in the CA cohort were notable for higher prevalence of cardiac tamponade (4.7% vs 0.5%, p<0.001) and vascular complications (2.6% vs 0.7%, p=0.016), which included vessel puncture or laceration, pseudoaneurysm/aneurysm, dissection, fistula, and hematoma formation (3.0% vs 0.6%, p=0.002). There were no significant differences between groups for pneumothorax, stroke, and DVT/PE (Figure 2 and Supplemental Table 2). Multivariable logistic regression demonstrated that any complication (cardiac tamponade, pneumothorax, stroke, vascular complication, hematoma, deep venous thrombosis and pulmonary embolism) was more likely to occur in the ablation cohort (OR 2.71, [95% CI 1.59, 4.53], p<0.001). Median length of stay for those undergoing VT ablation was longer (5 vs 4 days; p<0.001) and median cost of hospitalization was greater ($128,828 vs. $46,367; p <0.001). The majority of sarcoidosis patients in both groups were discharged home without assistance (87.9% vs. 82%; p=0.036).

**Figure 2.**
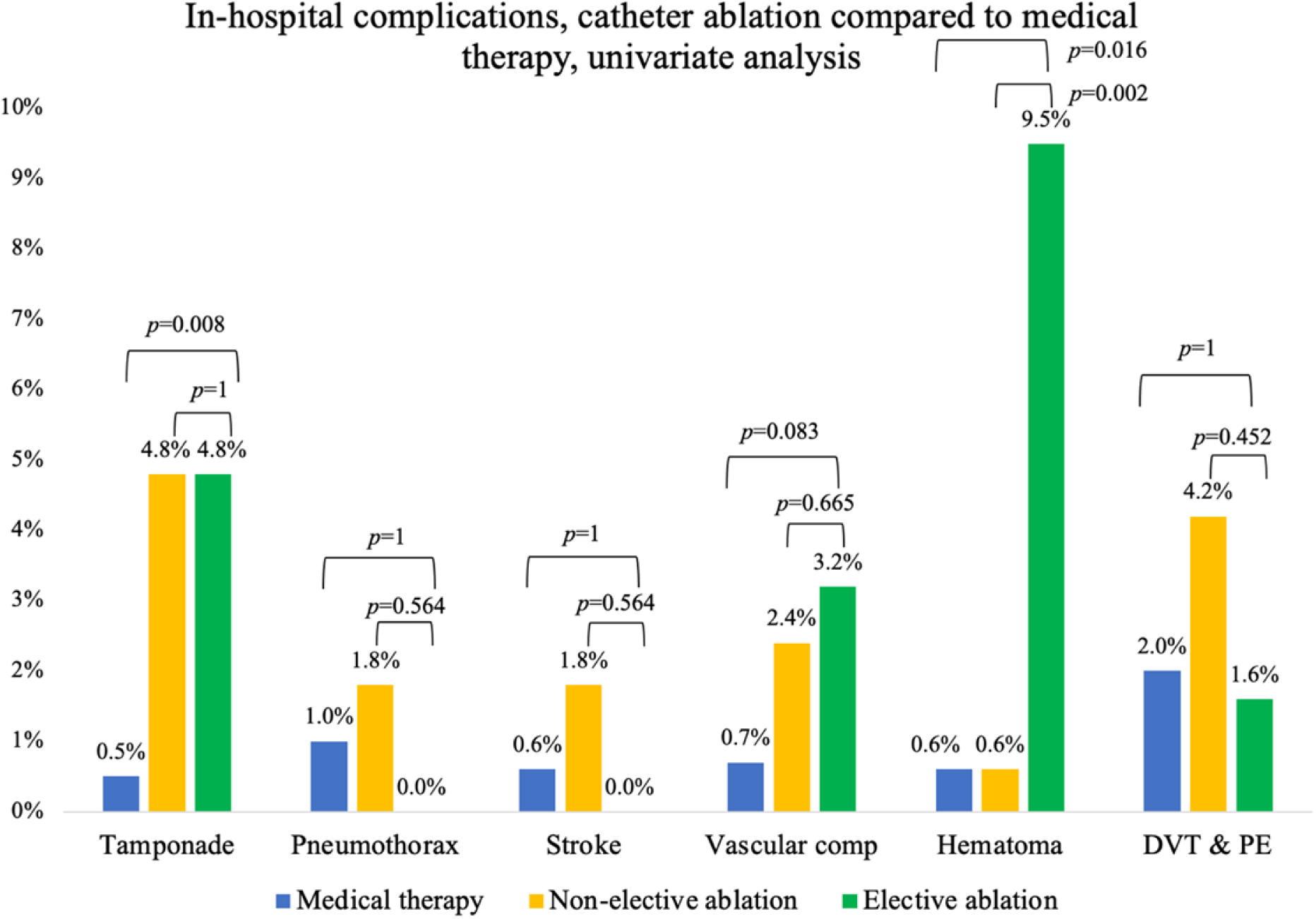
In-hospital complications (percentage) in patients with sarcoidosis undergoing VT ablation (index admission), univariate analysis. DVT=deep venous thrombosis, PE=pulmonary embolism, comp=complications

With respect to the index admission, univariate analysis showed that the use of mechanical circulatory support was higher in the ablation cohort (2.2% vs 0.3%, p=0.005). During index hospitalization, unadjusted analyses found that the composite of mortality, cardiac arrest, and cardiogenic shock trended toward significance in favor of ablation (7.4% vs 11.7%, p=0.067) (Table 2), however, this difference attenuated with multivariable analysis (OR 0.86, [95% CI 0.46, 1.51] p=0.603).

**Table 2.** In-hospital outcomes, medical therapy compared to catheter ablation, univariate analysis. Univariate analysis comparing in-hospital outcomes, including inpatient mortality, cardiac arrest, cardiogenic shock, mechanical circulatory support use between patients with sarcoidosis medically managed versus with catheter ablation. IQR=interquartile range, d=days

|  | Medical therapy<br>(N=1349) | Catheter ablation<br>(N=232) | P value |
| --- | --- | --- | --- |
| ***In-hospital outcomes*** |  |  |  |
| Mechanical circulatory support | 4 (0.3) | 5 (2.2) | 0.005 |
| Mortality | 35 (2.6) | 3 (1.3) | 0.350 |
| Cardiogenic shock | 56 (4.2) | 13 (5.6) | 0.409 |
| Cardiac arrest | 114 (8.5) | 4 (1.7) | 0.299 |
| Composite endpoint of mortality, cardiac arrest and cardiogenic shock | 158 (11.7) | 17 (7.4) | 0.067 |
| ***Hospitalization Characteristics*** |  |  |  |
| Median length of stay (IQR), d | 4.00 [2.00, 6.00] | 5.00 [3.00, 9.00] | <0.001 |
| Median cost of index hospitalization (IQR), \$ | 46367.00 [18248.00, 124576.00] | 128828.50 [75785.75, 237077.25] | <0.001 |
| ***Disposition at discharge*** |  |  |  |
| Home with no assistance | 1106 (82.0) | 203 (87.9) | 0.036 |
| Home with health aid | 123 (9.1) | 16 (6.9) | 0.337 |
| Transfer to subacute nursing or intermediate care facility | 83 (6.2) | 8 (3.5) | 0.142 |
| Other | 37 (2.7) | 4 (1.7) | 0.503 |

### Long-term outcomes

Unplanned 30, 90, and 180-day readmission outcomes were assessed between CA and medical therapy groups. Readmission rates were not significantly different in univariate analysis between medical therapy and catheter ablation at 30 days (10.8 vs 8.0%, p=0.266), 90 days (21.3 vs. 19.4%, p=0.606), or 180-days (30.7% vs. 27.0%, p=0.418) (Figure 3 and Supplemental Table 3). After multivariable adjustment, 30-day readmissions (OR 0.57, [95% CI 0.29,1.07] p=0.086) trended in favor of catheter ablation. After multivariable adjustment, catheter ablation was superior to medical therapy at 180-days (OR 0.28 [95% CI 0.12,0.65] *p*=0.003) whereas this trend was not observed with 90-day readmission rates (OR 0.69 [95% CI 0.37,1.31] p=0.255).

**Figure 3.**
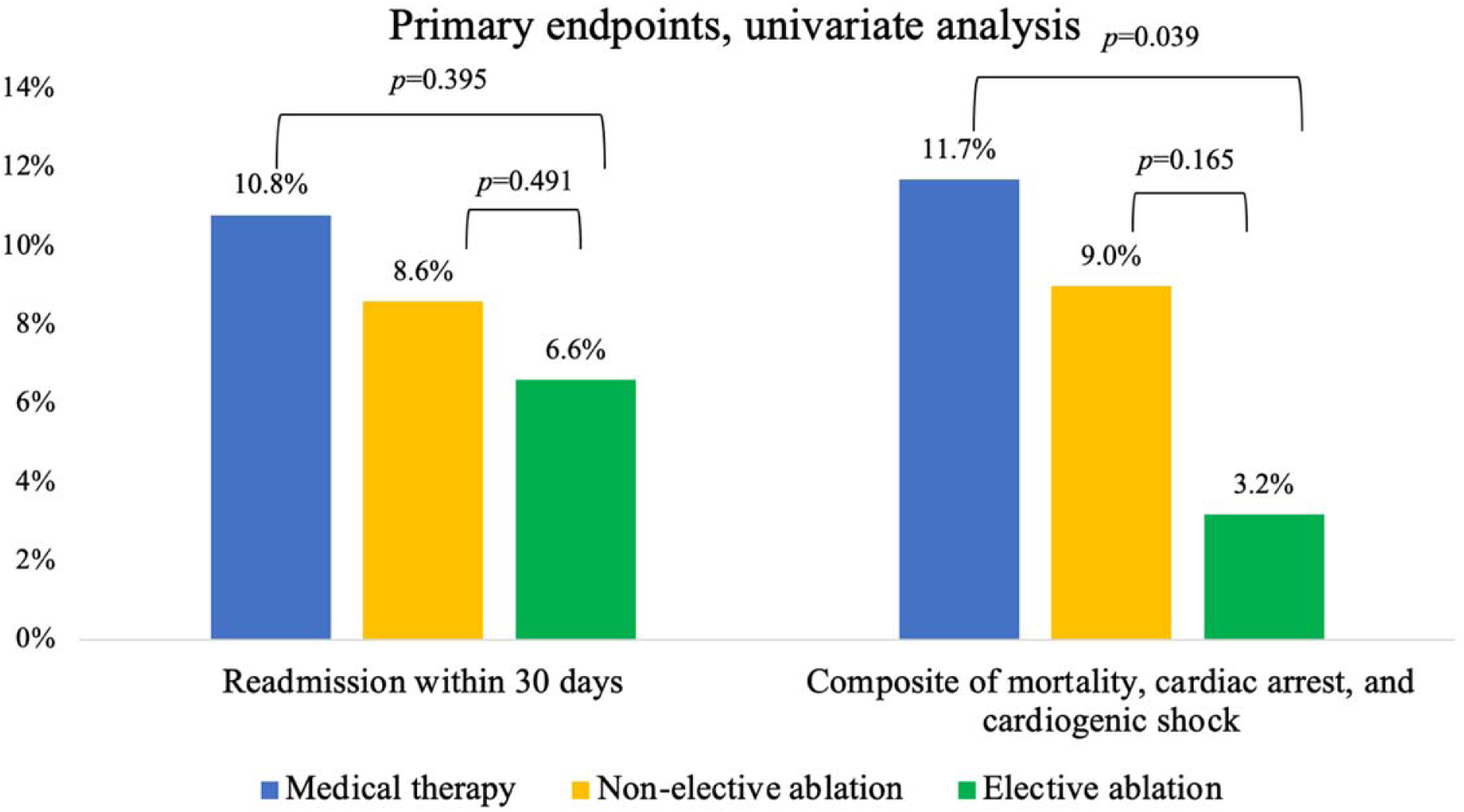
Comparison of readmission rates and in-hospital outcomes, including composite endpoint of inpatient mortality, cardiac arrest, cardiogenic shock between patients with sarcoidosis medically managed versus with catheter ablation, univariate analysis. Numbers in percentages.

Among patients who were readmitted within 30-days, the chief reason for readmission in univariate analysis were ventricular arrhythmias in both those undergoing CA and medical management (72.2% vs 40.1%; p=0.020). Other reasons for readmission included other arrhythmias, heart failure, device-related complications, pulmonary embolism, hypertension and coronary disease, vascular complications and hypotension. Heart failure (5.1% vs 0%; p=1), device-related complications (5.1% vs 0%; p=1), and hypertension/coronary disease (5.1% vs 0%; p=1) were more common reasons for readmission in the medical therapy group, whereas pulmonary embolism (5.6% vs 2.9% p=0.465), hypotension (5.6% vs 1.5% p*=*0.311), and vascular complications (11.1% vs. 0.7%; p*=*0.036) were more common in the catheter ablation group (Figure 4).

**Figure 4.**
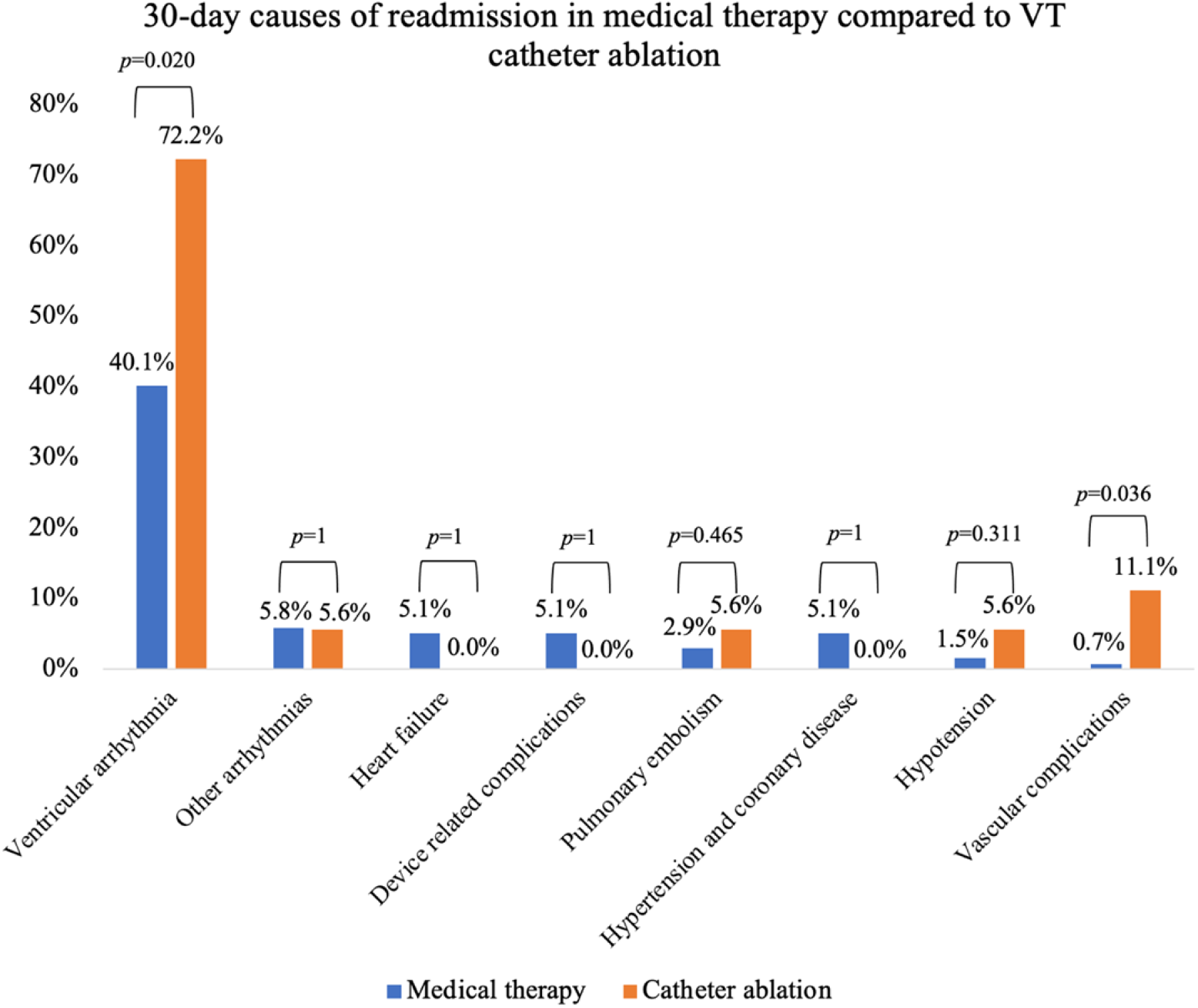
Common causes of readmission examined, stratified by cohort, medical therapy compared to catheter ablation. Numbers expressed as percentages of total readmissions.

### In-hospital and long-term outcomes, elective catheter ablation versus medical therapy

In univariate analysis, patients undergoing elective CA for VT experienced a significant improvement in composite outcome of mortality, cardiac arrest, and cardiogenic shock (3.2% vs. 11.7%, p=0.039) compared to medical therapy patients (Table 3 and Figure 3). Patients undergoing elective CA for VT were more likely to have an ICD compared to those medically managed alone (66.7% v 41.4%; *p*=<0.001) (Table 3). Univariate analyses of patients undergoing non-elective VT ablation did not have similar benefit versus medical therapy (9.0% vs 11.7%, p=0.359) (Supplemental Table 4). After multivariable adjustment, elective VT ablation had no clear benefit over medical therapy in the primary composite endpoint (OR 0.94 [95% CI 0.13,4.27] p=0.940). Further, all patients who underwent elective ablation went home with no assistance (100.0% vs. 82.0%, p<0.001) compared to those medically managed (Table 3). While univariate analysis did not show a significant difference in 30-day readmission rates between medical therapy and elective catheter ablation (6.6% vs. 10.8%, p=0.395), multivariable logistic regression demonstrated that patients with elective CA were less likely to be readmitted after 30 days (OR 0.23 [95% CI 0.05,0.90] p=0.042).

**Table 3.** In-hospital outcomes, medical therapy compared to elective catheter ablation, univariate analysis. Univariate analysis comparing in-hospital outcomes, including inpatient mortality, cardiac arrest, cardiogenic shock, mechanical circulatory support use between patients with sarcoidosis medically managed versus with elective catheter ablation. IQR=interquartile range, d=days

|  | **Medical therapy**<br>**(N=1349)** | **Elective ablation**<br>**(N=63)** | **P value** |
| --- | --- | --- | --- |
| ***In-hospital outcomes*** |  |  |  |
| Mechanical circulatory support | 4 (0.3) | 3 (4.8) | 0.003 |
| Mortality | 35 (2.6) | 0 (0.0) | 0.401 |
| Cardiogenic shock | 56 (4.2) | 2 (3.2) | 1 |
| Cardiac arrest | 114 (8.5) | 0 (0.0) | 0.008 |
| Composite endpoint of mortality, cardiac arrest and cardiogenic shock | 158 (11.7) | 2 (3.2) | 0.039 |
| ***Device and drug therapy*** |  |  |  |
| Implantable cardioverter-defibrillator | 558 (41.4) | 42 (66.7) | <0.001 |
| Immunosuppression and device therapy | 147 (10.9) | 7 (11.1) | 1 |
| ***Hospitalization characteristics*** |  |  |  |
| Median length of stay (IQR), d | 4.00 [2.00, 6.00] | 3.00 [2.00, 5.00] | 0.36 |
| Median cost of index hospitalization (IQR), \$ | 46367.00<br>[18248.00, 124576.00] | 121596.00<br>[73860.50, 188144.00] | <0.001 |
| ***Disposition at discharge*** |  |  |  |
| Home with no assistance | 1106 (82.0) | 63 (100.0) | <0.001 |
| Home with health aid | 123 (9.1) | 0 (0.0) | 0.005 |
| Transfer to subacute nursing or intermediate care facility | 83 (6.2) | 0 (0.0) | 0.048 |
| Other | 37 (2.7) | 0 (0.0) | 0.407 |

**Table 4.** In-hospital outcomes, elective ablation compared to non-elective ablation, univariate analysis. Univariate analysis comparing in-hospital outcomes, including inpatient mortality, cardiac arrest, cardiogenic shock, mechanical circulatory support use between patients with sarcoidosis undergoing elective versus non-elective catheter ablation.

|  | **Elective ablation**<br>**(N=63)** | **Non-elective ablation**<br>**(N=168)** | **P value** |
| --- | --- | --- | --- |
| ***In-hospital outcomes*** |  |  |  |
| Mechanical circulatory support | 3 (4.8) | 2 (1.2) | 0.127 |
| Mortality | 0 (0.0) | 3 (1.8) | 0.564 |
| Cardiogenic shock | 2 (3.2) | 11 (6.5) | 0.523 |
| Cardiac arrest | 0 (0.0) | 4 (2.4) | 0.577 |
| Composite endpoint of mortality, cardiac arrest and cardiogenic shock | 2 (3.2) | 15 (9.0) | 0.359 |
| ***Device and drug therapy*** |  |  |  |
| Implantable cardioverter-defibrillator | 42 (66.7) | 80 (47.6) | 0.015 |
| Immunosuppression and device therapy | 7 (11.1) | 28 (16.7) | 0.399 |
| ***Hospitalization characteristics*** |  |  |  |
| Median length of stay (IQR), d | 3.00 [2.00, 5.00] | 6.00 [4.00, 10.25] | <0.001 |
| Median cost of index hospitalization (IQR), \$ | 121596.00 [73860.50, 188144.00] | 132528.00 [78263.75, 263141.50] | 0.131 |
| ***Disposition at discharge*** |  |  |  |
| Home with no assistance | 63 (100.0) | 139 (83.2) | <0.001 |
| Home with health aid | 0 (0.0) | 16 (9.6) | 0.007 |
| Transfer to subacute nursing or intermediate care facility | 0 (0.0) | 8 (4.8) | 0.111 |
| Other | 0 (0.0) | 4 (2.4) | 0.577 |

### In-hospital and long-term outcomes, elective ablation compared to non-elective ablation

Baseline characteristics between elective and non-elective catheter ablation groups were overall similar except for higher prevalence of implantable cardioverter-defibrillators in the elective group (66.7% vs. 47.6%) (Supplemental Table 5). Compared to non-elective ablation, patients undergoing elective VT CA patients were more likely to have shorter length of stay, (3 days vs 6 days; p<0.001) and go home with no assistance (100% vs 83.2%; p<0.001). There were no significant differences in complications with respect to tamponade, pneumothorax, stroke, or DVT/PE. There was an increase in hematomas in the elective CA group compared to the non-elective CA group (0.6% vs. 9.5%, p=0.002) (Figure 2).

When comparing elective VT CA to non-elective VT CA, univariate analysis found no significant difference, but a trend towards improvement in readmission rates, MCS, mortality, cardiogenic shock, or cardiac arrest. Multivariable analysis demonstrated a trend towards improvement in the composite endpoint with elective CA (OR 0.52 [95% CI 0.27,1.01] p=0.051). Multivariable regression was not performed for readmission data between the two groups as the number of events was too small to adjust for the covariates of interest. Within the catheter ablation cohort, ventricular arrhythmias were a more common cause of readmission in the non-elective group compared to elective ablation (86% vs. 25%; p=0.028) (Figure 5).

**Figure 5.**
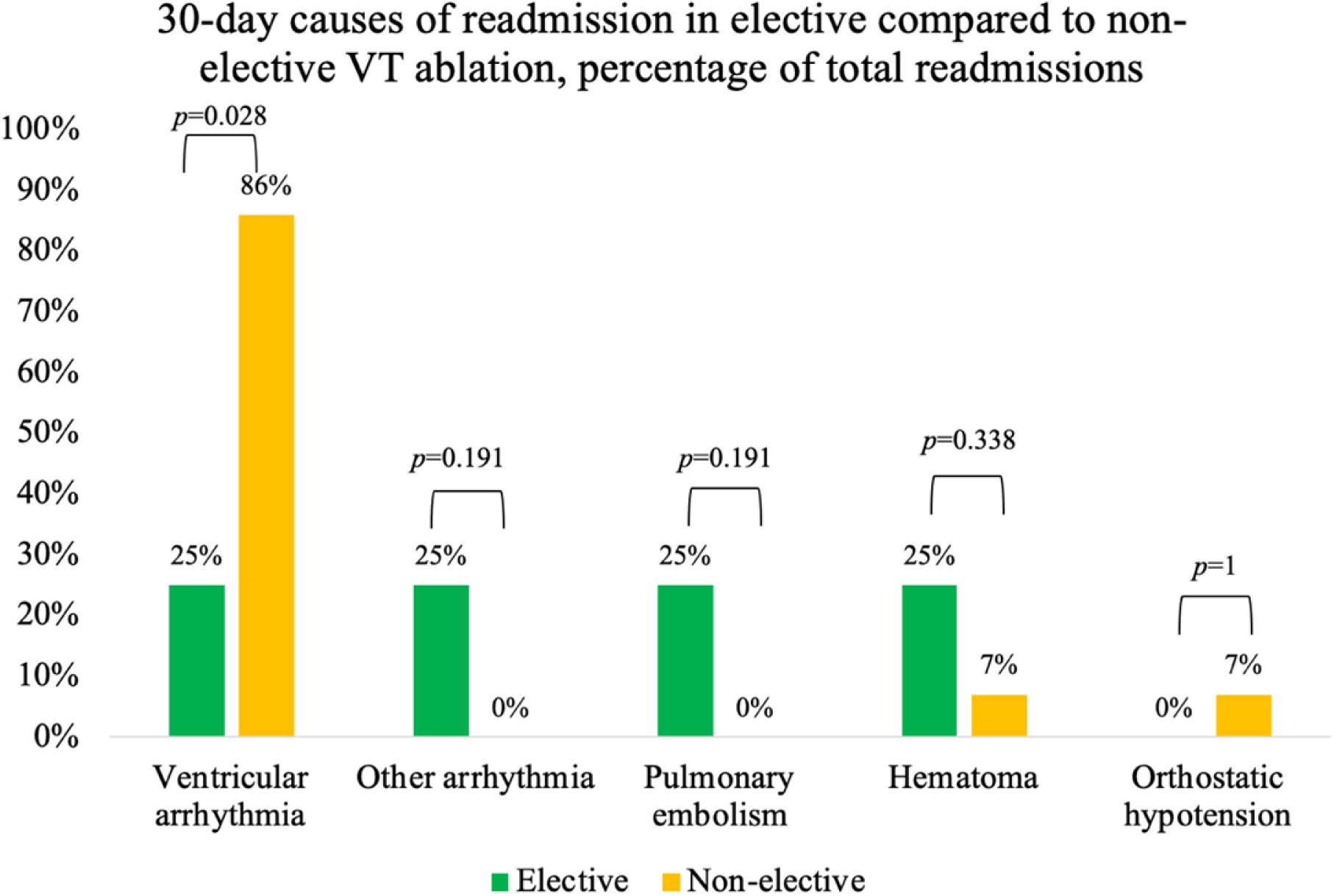
Causes of readmission in sarcoidosis patients undergoing elective compared to non- elective catheter ablation. Numbers expressed as percentages of total readmissions.

## Discussion

There are few real-world studies evaluating in-hospital and short-term readmissions outcomes in the sarcoidosis population undergoing VT ablation as compared to medical therapy. To date, this is the largest cohort study to study VT ablation versus medical therapy in the sarcoidosis population. There were a number of unique findings in this study. First, sarcoidosis patients undergoing VT ablation trended towards a lower composite of in-hospital mortality, cardiogenic shock, and cardiac arrest compared to those who were medically managed during the index hospitalization when catheter ablation took place. In univariate analysis, elective catheter ablation was superior to medical therapy in the primary composite endpoint. After multivariable adjustment, elective ablation demonstrated a strong trend over non-elective catheter ablation in the composite endpoint of in-hospital mortality, cardiogenic shock and cardiac arrest. Second, there was no increase in 30-day readmission rates for patients with sarcoidosis treated with VT ablation, compared to medical therapy. After multivariable adjustment, patients undergoing elective catheter ablation were less likely to be readmitted 30-days after VT ablation. Third, complications in both elective and non-elective VT ablation as well as mechanical circulatory support use remained low. These results in total support the safety of VT ablation when compared to medical therapy in patients with sarcoidosis, and in particular, elective catheter ablation.

An additional important finding in this study is the unadjusted analysis demonstrating a reduction in the composite endpoint of mortality, cardiac arrest, and cardiogenic shock among patients who underwent elective VT ablation compared to those who were medically managed. After risk factor adjustment, this finding was no longer statistically significant. Elective ablation was also associated with shorter length of stay and hospital discharge home without assistance. Multivariable analysis also demonstrated that patients with elective CA were less likely to be readmitted within 30 days. One explanation may be that elective ablation patients are inherently healthier and therefore are more likely to avoid in-hospital cardiac mortality, cardiac arrest and shock secondary to prolonged VT ablation procedures that can promote hemodynamic instability. Additionally, VT ablation is complex, with outcomes that are improved with periprocedural planning including the use of advanced imaging modalities. Computed tomography and cardiac MRI, which is not readily available during inpatient admission at many centers, may aid in a critical way for procedural success by providing three dimensional maps that help identify anatomic corridors and areas of scar. Third, patients undergoing elective ablation may have been treated with immunosuppressants and represent a different stage of disease compared to those who undergo non-elective ablation. Kaur et al demonstrated that outcomes of VT ablation are worse in the inflammatory phase of sarcoidosis compared with the scar phase. In 24 patients presenting with drug-refractory VT, acute procedural success, defined as non-inducibility of the clinical VT, was attained in 10 out of 17 patients with inflammation compared with complete success in 6 out of 7 without inflammation. Over a mean period of 5.7 years, VT recurrence was 58.8% in those with ongoing inflammation compared to 14.2% with scar.^2^ In another observational study assessing determinants of long-term outcomes in 158 CS patients undergoing catheter ablation for VT, inflammation on FDG-PET was associated with worse long-term prognosis, including recurrent VT, death, or need for heart transplantation.^6^ These results suggest inflammation should be treated, if possible, prior to catheter ablation.

Alternatively, it is important to acknowledge that some of the factors that were adjusted for in the first multivariate analysis may not be permanent patient characteristics. Heart failure, renal failure, diabetes, liver disease, chronic obstructive pulmonary disease, anemia, electrolyte and coagulopathies can be modified and optimized. Additionally, VT storm necessitating admission may be the causative force worsening heart failure and other organ dysfunction. If so, these findings coupled with the high recurrence rate of VT treated with medical therapy would argue for elective VT ablation in the CS population. Further research is warranted to determine if there is a subgroup of CS patients admitted with VT who are better served with an initial conservative management strategy coupled with risk factor optimization followed by elective VT ablation.

Limited cohort studies with small numbers of patients have shown that catheter ablation reduces VT recurrence in cardiac sarcoidosis patients. Further, VT ablation for sarcoidosis patients is effective in reducing arrhythmia burden and allows for de-escalation of antiarrhythmic therapy, the side effects of which also carry additional risk.^13^ In a single center study, out of nine CS patients undergoing VT ablation, five had no VT recurrence and the other four had a substantial reduction in VT episodes at 3 months. In the same study, nearly 50% of the patients could not attain arrhythmia control with medical therapy alone.^8^ Another group demonstrated an arrhythmia free survival in 40% of patients and a reduction in arrhythmia burden in 90% of 31 CS patients undergoing VT ablation.^5^ Additionally, mortality has been low in VT catheter ablation procedures. According to a systematic review on CA for VT in patients with CS, procedural mortality was 0%^17^, consistent with the low inpatient mortality observed in sarcoidosis patients in this study.

While this data has been promising, there is little data to support superiority of CA over medical therapy in VT secondary to cardiac sarcoidosis. Analysis from the National Inpatient Sample compared outcomes in patients who were medically managed to catheter ablation and showed a trend, but failed to show a statistically significant reduction in mortality with ablation.^18^ Our analyses are consistent with these studies, but in addition demonstrated for the first time a reduction in the combined endpoint of death, cardiogenic shock and cardiac arrest in patients undergoing elective VT ablation.

The observed in-hospital complications in this study are similar to published data in patients with structural heart disease undergoing VT ablation. In a large single center study of 548 patients with structural heart disease and a small percentage with idiopathic VT undergoing 722 ablation procedures, there was a 6.2% major complications rate, with access site vascular complications being most common at 3.6%; cardiac tamponade was only prevalent in 0.4% of cases.^19^ In another large study of 528 patients undergoing CA for VT with structural heart disease, the rate of post procedural tamponade was 2%.^14^ An additional NRD study in MI-associated VT ablation, the rate was 2.6%.^20^ In this study, the rate of cardiac tamponade at 4.7% is slightly higher than published literature. This is likely due to other competing factors that we could not account for including operator experience, procedure time, and access site including need for epicardial access. Given the proclivity for sarcoidosis to have patchy involvement of the myocardium, epicardial access for VT ablation is frequently necessary in cases when the VT cannot be ablated using an endocardial approach.^8, 9, 13, 17^ Although tamponade was the most common complication in the elective CA group, other feared complications such as stroke, pneumothorax, vascular complications, and DVT/PE were low. It is unclear why hematomas were more common in the elective VT ablation group. Perhaps these patients were more likely to be on anticoagulation during the periprocedural period and thus more susceptible to bleeding.

The primary cause for readmission in this study was ventricular arrhythmia; however, of the patients undergoing CA, VAs were far less common in the elective CA group. The difference between elective and non-elective CA may be explained by several factors including transient breakthrough arrhythmia, abrupt de-escalation of anti-arrhythmic medications, or procedural failure. Additionally, patients in the elective group may have had PET-CT imaging available, either showing improvement or resolution of inflammation prior to scheduling ablation, while patients in the non-elective group may not have been able to obtain advance inpatient PET-CT imaging, thereby making treatment less effective if active inflammation were present. Patients presenting with VAs may also be more frequently readmitted due to shocks from ICDs, which provide life-saving therapies and prompt further medical attention. Other causes of readmission including non-VA arrhythmias, device-related complications, and coronary disease remained low and there was no signal of increased harm either during index admission or on readmission. As anticipated, vascular complications were higher in the CA group. Despite the VT CA cohort having greater baseline prevalence of heart failure, there were zero 30-day readmissions due to heart failure following CA for VT, suggesting that CA may be crucial in preventing short-term heart failure readmissions. Further studies are warranted to assess the potential benefit of ablation over medical therapy to prevent heart failure.

Growing evidence suggests that timing is an important element of VT ablation in patients with CS. Studies suggest that VT ablation in the scar phase of the disease, after active inflammation has been adequately treated, is associated with the greatest freedom from VT recurrence post-ablation.^2, 6^ In-line with these findings, recent expert consensus recommends a stepwise approach to the management of VT in CS, with intensification of immunosuppression and antiarrhythmic drug therapy preceding catheter ablation in the absence of urgent indications for the latter (e.g. VT storm, ventricular fibrillation refractory to medical management).^6, 7^ Alternatively, new evidence not specific to cardiac sarcoidosis, has emerged in favor of earlier VT ablation after first ICD shock or as an upfront therapy prior to escalation of antiarrhythmics.^15, 16, 21^ Most recently, an observational study comparing early VT ablation for VT storm in 58 subjects compared to medical therapy in 71 subjects proved ablation superior in reduction of VT recurrence, storm recurrence, iatrogenic complications, cardiovascular hospitalizations, and cumulative days in hospital in follow up.^22^ While the role of inflammation in cardiac sarcoidosis is unique, this growing body of evidence for early intervention in VT combined with our findings that elective ablation in CS reduces cardiogenic shock, cardiac arrest, and inpatient mortality combined, may lead to a paradigm shift to earlier VT ablation, possibly while inflammation is not completely resolved. Patients with sarcoidosis and recurrent VT may not have time to wait until inflammation has been treated due to the high morbidity and mortality of this disease. VT ablation may obviate the need for long-term immunosuppressants or antiarrhythmics. Importantly, our findings indicate that VT ablation can be performed with favorable short-term outcomes and low rates of complications.

Limitations of this study are primarily related to characteristics of the NRD. As part of a large administrative database relying on billing codes, NRD entries are susceptible to misclassification and their analysis subject to residual confounding due to non-randomized treatment assignment. The temporality of outcomes such as cardiac arrest and cardiogenic shock relative to the timing of catheter ablation could not be established. There is also a limit to the database’s granularity as it lacks data on procedural details, operator experience, pharmacotherapies, peri-procedural imaging and laboratory values. Patient level data including cardiac MRI or FDG-PET findings, which may be crucial in procedural planning and understanding of VT substrate, were not available. We were also unable to provide procedure level characteristics such as epicardial ablation and whether there was absence of VT inducibility, which may suggest lower recurrence rates.^23^ Selection bias may significantly affect the benefit seen in patients undergoing elective VT ablation as this may represent a healthier patient population that would have less risk of adverse outcomes compared to those who were admitted non-electively. Operators may have selected healthier patients to undergo VT ablation rather than medical therapy. There may also be a selection bias in patients with devices; having a defibrillator may portend better outcomes. Further, management of patients with VT, and in particular catheter ablation, took place at large, metropolitan teaching hospitals. At index admission, hospitalization requiring VT ablation was costlier than admission without VT CA. Further analyses to determine long term costs beyond the index admission should be pursued. Additionally, the NRD does not capture out-of-hospital death and hence our analysis could not account for the competing risk for post-discharge death and readmission. Lastly, the NRD does not include follow-up data for greater than 1 year, hence longer-term endpoints could not be investigated.

## Conclusion

In a national database of patients admitted with sarcoidosis and ventricular tachycardia, when compared to medical therapy, catheter ablation resulted in similar 30-day readmission rates with a trend towards reduction in the composite endpoint of inpatient mortality, cardiogenic shock, and cardiac arrest and a significant reduction in patients undergoing elective VT ablation. Procedure related complications were low. Elective VT ablation patients had lower rates of 30- day readmission and the combined endpoint of inpatient mortality, cardiogenic shock or cardiac arrest, although this may be due to a lower risk clinical profile or pre-procedural optimization. Despite data from previous publications suggesting worse outcomes compared to other VT ablation subtypes, our findings support CA, ideally electively, as a possible alternative to medical therapy alone.

## Data Availability

The Nationwide Readmissions Database is publicly available data set sponsored by the Agency for Healthcare Research and Quality as part of the Healthcare Cost and Utilization Project.

https://hcup-us.ahrq.gov/nrdoverview.jsp

